# SARS-COV-2 comorbidity network and outcome in hospitalized patients in Crema, Italy

**DOI:** 10.1101/2020.04.14.20053090

**Authors:** Gianpaolo Benelli, Elisabetta Buscarini, Ciro Canetta, Giuseppe La Piana, Guido Merli, Alessandro Scartabellati, Giovanni Viganò, Roberto Sfogliarini, Giovanni Melilli, Roberto Assandri, Daniele Cazzato, Davide Sebastiano Rossi, Susanna Usai, Guido Caldarelli, Tommaso Gili, Irene Tramacere, Germano Pellegata, Giuseppe Lauria

**Author notes:** Corresponding author Prof. Giuseppe Lauria, Department of Clinical Neuroscience, IRCCS Foundation “Carlo Besta” Neurological Institute, Via Celoria 11 - 20133, Milan, Italy. Contributed equally (Gianpaolo Benelli, Elisabetta Buscarini, Ciro Canetta, Giuseppe La Piana, Guido Merli, Alessandro Scartabellati, Giovanni Viganò).

## Abstract

No systematic data on hospitalized SARS-COV-2 patients from Western countries are available. We report onset, course, correlations with comorbidities, and diagnostic accuracy of nasopharyngeal swab in 539 individuals suspected to carry SARS-COV-2 admitted to the hospital of Crema, Italy. All individuals underwent clinical and laboratory exams, SARS-COV-2 reverse transcriptase-polymerase chain reaction on nasopharyngeal swab, and chest X-ray and/or computed tomography (CT). Data on onset, course, comorbidities, number of drugs including angiotensin converting enzyme (ACE) inhibitors and angiotensin-II-receptor antagonists (sartans), follow-up swab, pharmacological treatments, non-invasive respiratory support, ICU admission, and deaths were recorded. Among 411 SARS-COV-2 patients (66.6% males) median age was 70.5 years (range 1-99). Chest CT was performed in 317 (77.2%) and showed interstitial pneumonia in 304 (96%). Fatality rate was 17.5% (74% males), with 6.6% in 60-69 years old, 21.1% in 70-79 years old, 38.8% in 80-89 years old, and 83.3% above 90 years. No death occurred below 60 years. Non-invasive respiratory support rate was 27.2% and ICU admission 6.8%. Older age, cough and dyspnea at onset, hypertension, cardiovascular diseases, diabetes, renal insufficiency, >7 drugs intake and positive X-ray, low lymphocyte count, high C-reactive protein, aspartate aminotransferase and lactate dehydrogenase values, and low PO_2_ partial pressure with high lactate at arterial blood gas analysis at admission were significantly associated with death. Use of ACE inhibitors or sartans was not associated with outcomes. Comorbidity network analysis revealed homogenous distribution of deceased and 60-80 aged SARS-COV-2 patients across diseases. Among 128 swab negative patients at admission (63.3% males) median age was 67.7 years (range 1-98). Chest CT was performed in 87 (68%) and showed interstitial pneumonia in 76 (87.3%). Follow-up swab turned positive in 13 of 32 patients. Using chest CT at admission as gold standard on the entire study population of 539 patients, nasopharyngeal swab had 80% sensitivity. SARS-CoV-2 caused high mortality among patients older than 60 years and correlated with pre-existing multiorgan impairment. ACE inhibitors and sartans did not influence patients’ outcome.

## Introduction

Since December 2019 outbreak in China of the novel coronavirus infection, designated SARS-CoV-2 and termed Covid-19, the disease has quickly overflowed worldwide, to date mostly affecting the European countries and the United States. SARS-CoV-2 causes a clinical syndrome encompassing asymptomatic or oligosymptomatic flu-like course, gastrointestinal disturbances, mild pneumonia, acute respiratory distress and death.^1-3^

After the first Italian patient was notified on February 18^th^, 2020 at the hospital of Codogno, 25 km far from Crema, the total number of SARS-CoV-2 patients exponentially increased in the area. The municipality of Crema, with about 34,000 inhabitants, experienced a rapid overload of the healthcare structures due to a massive influx of suspected SARS-CoV-2 patients.

We sought to perform a systematic analysis of hospitalized SARS-CoV-2 patients soon after the outbreak and outline possible correlations with comorbidities and drug intake. To this aim, we investigated all consecutive individuals suspected to harbor SARS-CoV-2 and admitted to the General Hospital of Crema between February 21^st^ and March 13^rd^, 2020 and followed-up until March 19^th^, 2020. We provided data on onset, clinical history, laboratory and radiological findings, course of the disease, age and sex-stratified fatality rate, and diagnostic accuracy of the nasopharyngeal swab. We applied a network analysis to assess the interactions between comorbidities across age groups and outcomes.

We trust our findings could provide the global community and healthcare systems with useful information to better face the current health emergency and identify novel strategies to prevent worst outcomes in patients.

## Methods

On February 19^th^, 2020 the General and Health Directorate of the Hospital of Crema met to update and carry out the available procedures to cope with the hospitalization of patients with a potential viral spread, based on 2009 SARS and H1N1 pandemic strategic plan revised on December 2014 after Ebola outbreak. Starting on February 21^st^, 2020 the Emergency Department set up a triage for any individual either reporting or presenting with fever (above 37.5°C), cough or dyspnea, or having had contact with SARS-CoV-2 carriers. All individuals admitted to the hospital underwent body temperature and pulse oximetry (SO_2_) recording, hematological screening, chest X-ray and/or computed tomography (CT) and nasopharyngeal swab. Swabs were stored at +4°C and immediately shipped to one of the laboratory of virology accredited by the Lombardy Region for diagnostic SARS-COV-2 real-time polymerase chain reaction assay. Based on clinical, laboratory and radiological findings, patients were discharged to home in quarantine or hospitalized.

Demographic data, date of onset and type of symptoms, comorbidities (e.g. hypertension, cardiovascular disorders, diabetes, pulmonary diseases, active and previous malignancies, renal insufficiency and any other known condition), current pharmacological treatments, use of angiotensin converting enzyme inhibitors (ACE inhibitors) and angiotensin-II-receptor antagonists (sartans), and number of drugs were recorded. All available clinical data including hematological and radiological exams, treatments, respiratory support with continuous positive airway pressure (CPAP) or non-invasive ventilation (NIV), ICU admission and deaths were recorded. Chest X-ray and CT scan were scored as positive or negative based on radiologists’ written report. Acute reticular pattern at X-ray and the presence of single or multiple ground-glass and/or consolidative lung opacities were considered indicative of interstitial pneumonia (X-ray and CT positive). Pleural and pericardial effusion, and lymphadenopathy at CT scan were also recorded.

The hospital of Crema is fully equipped with a computerized recording system that generates a unique code for any visit and exam. All patients were anonymized and locked to the unique code assigned at the admission, and all data were included in an electronic database. The Institutional Review Board of the Hospital of Crema has approved the study. All the experiment protocol for involving human data was accordance to guidelines of national/international/institutional or Declaration of Helsinki in the manuscript. Written informed consent was obtained from all participants and legally authorized representatives of minors below 18 patients and dead patients gave consent for data that could be used for clinical studies.

### Comorbidity network analysis

Two different sets of nodes were considered for the network analysis: 252 SARS-Cov-2 positive patients and 13 possible diseases (cardiovascular disease, respiratory diseases, diabetes, hypertension, renal insufficiency, cancer, arthritis, obesity, asthma, obstructive sleep apneas, dyslipidemia, chronic obstructive pulmonary disease, atrial fibirllation) they could suffer for. A rectangular matrix patients (rows) vs diseases (columns) (PvsD) was obtained by associating to the entry PvsD(i,j) one or zero according to the fact that patient i had been suffering for the disease j or not. The information in this matrix can be use to define an adjacency matrix ^4^ representing a graph, where two different sets of nodes were included (patients and diseases) and for each non-zero entry of the matrix a link between two nodes was included. The resulting graph is known as a bipartite network ^5^ that is, a graph where nodes can be decomposed into two disjoint sets A and B such that edges connect only elements of one set with elements of the other set. A common practice for bipartite data mining is to find a projection of the whole graph over one of the two sets. The A-projection is composed of a network containing only A-nodes, where two A-nodes are connected when they have at least N common neighboring B-node ^5^.

Comorbidity networks are typical bipartite where one set is given by the patients and the other by their diseases ^6^. In this study, it was obtained by firstly dividing the cohort of patients in three classes of age: 40-60 years, 61-80 years, and 81-100 years. The networks have been calculated as the projection of the patient-disease bipartite network on the diseases space. This means that two diseases are connected by one edge if at least one patient suffer from both and the weight of this edge is given by the total number of patients in this situation. Such final monopartite disease networks have been filtered by fixing N to the 10% of the number of patients included in each class. The three comorbidity networks have been analyzed in terms of nodes weighted degree ^4^, that measures a node’s total level of involvement in the network, and of modules detection ^7^ which iteratively optimizes local communities until global modularity can no longer be improved given perturbations to the current community state. Thereafter, the same cohort has been divided in deceased and survived patients. Comorbidity networks were obtained for the two groups in the same way described above and the same topological analysis has been done.

In order to test the statistically significance of the difference between deceased and survived networks, we decided to compare their normalized Laplacian matrices. The normalized Laplacian matrix is a transformation of the adjacency matrix, whose eigenvalues describe aspects related to the global network structure and dynamic interactions among network parts ^8^. We derived, as reported in Supplementary Materials, the null model of the difference of the two normalized Laplacian matrices and calculated the p-value associated.

## Statistical analysis

Descriptive statistics were provided in terms of absolute number and percentage for categorical data, and mean with standard deviation (SD) and median with value range for continuous data. Differences between groups were assessed by the chi-square test or Mann-Whitney test, as appropriate; Bonferroni correction was used to adjust for multiple testing. Sensitivity and specificity with the corresponding 95% confidence interval (95% CI), and positive and negative predicting values of the nasopharyngeal swab were calculated using as reference the CT scan.

Comorbidity network has been analyzed by dividing the patients in three classes of age and outcome (deceased and survived) at last day of follow-up. For each group, the comorbidity network was derived according to the constraint that at least 10% of patients must share two diseases.

### Data availability and sharing

The data that support the findings of this study are available from the corresponding author.

## Results

### Clinical features of the study population

We collected data from 539 consecutive patients admitted to the hospital. All underwent nasopharyngeal swab, whose result was available within 24 hours on average. Overall, 411 patients were swab positive and 128 negative. SARS-CoV-2 patients reported the onset of symptoms for 5.3±4.3 days, whereas swab negative patients for 5.7±4.1 days. The frequency of fever, cough and dyspnea did not differ between groups. Table 1 reports the prevalence of the symptoms at onset.

**Table 1.** Way of arrival at the hospital and symptoms reported by SARS-CoV-2 and swab negative patients.

| n (%) | Positive<br>(411) | Negative<br>(128) |
| --- | --- | --- |
| **Arrival at hospital** |  |  |
| *Ambulance* | 196 (47.7) | 36 (28.1) |
| *Own means* | 215 (52.3) | 92 (71.9) |
| **Fever** | 346 (84.2) | 97 (75.8) |
| **Cough** | 151 (36.7) | 44 (34.4) |
| **Dyspnoea** | 131 (31.9) | 34 (26.6) |
| **Syncope** | 41 (9.9) | 6 (4.7) |
| **Fall with trauma** | 23 (5.6) | 7 (5.4) |
| **Nausea** | 18 (4.4) | 5 (3.9) |
| **Vomit** | 16 (3.9) | 8 (3.7) |
| **Diarrhoea** | 15 (3.6) | 5 (3.9) |
| **Confusion/psychomotor restlessness** | 15 (3.6) | 6 (4.7) |
| **Headache** | 10 (2.4) | 2 (1.6) |
| **Coma** | 9 (2.2) | 4 (3.1) |
| **Diffuse myalgia/arthralgia** | 7 (1.7) | 2 (1.6) |
| **Thoracic pain** | 7 (1.7) | 10 (7.8) |
| **Abdominal pain** | 5 (1.2) | 2 (0.9) |
| **Pharyngodinia** | 4 (1.0) | 7 (5.4) |
| **Recent pneumonia in treatment** | 4 (1.0) | 3 (2.3) |

Thirteen patients arrived in coma. Of them, 9 were SARS-CoV-2 patients and 5 died. The other 4 patients were negative at nasopharyngeal swab; 3 died and 1 was hospitalized. Another 21 patients presented with disorientation and/or with psychomotor restlessness; 15 were SARS-CoV-2 patients and 2 died. The other 6 patients were swab negative and 4 died. Notably, 41 (9.9%) SARS-CoV-2 patients reported or had at arrival one or more syncope; 14 of them had a fall with trauma and 5 died. Another 6 (4.7%) swab negative patients reported syncope and 1 died.

At admission, body temperature was slight above 37°C both in SARS-CoV-2 and swab negative patients. Nevertheless, the large majority of patients reported the intake of paracetamol during the previous hours, thus influencing the assessment at arrival. Similarly, the mean value of pulse oximetry in patients who arrived in ambulance was influenced by the early treatment with oxygen mask during transportation. The mean value of those who reached the hospital by their own means did not differ between SARS-CoV-2 (93%, range 60-100) and swab negative (92%, range 50-100) patients.

The rapidly increasing number of patients needing respiratory support required a fast reorganization of the entire hospital. In particular, the number of sub-intensive care beds to assist patients with CPAP and NIV was increased by 324% and that of ICU beds by 100% (fig. 1).

**Figure 1.**
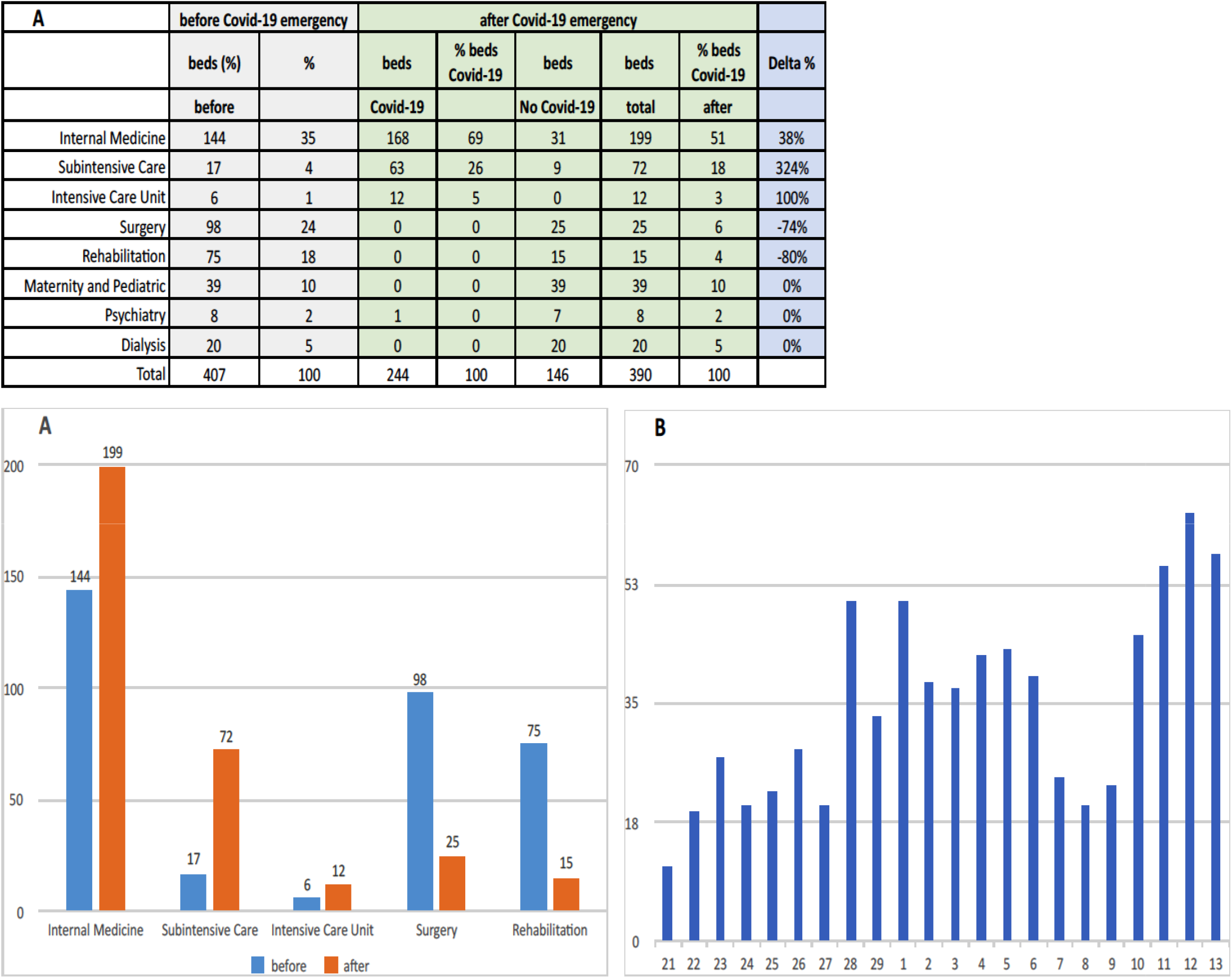
Distribution of beds before and after Covid-19 emergency **(A)** and daily number of access to the Emergency Department **(B)** between February 21 and March 13, 2020

### Course and outcome in SARS-CoV-2 patients

Among 411 in SARS-CoV-2 patients at nasopharyngeal swab, the median age was 70.5 years (range 1-99), with a preponderance of males (66.6%). One 87 year-old had confusion, psychomotor restlessness, vomit and fever; he arrived in coma with 38.5°C of body temperature and stiff neck, and died after 3 days with diagnosis of possible meningoencephalitis. Of 411 patients, 262 (63.7%) were hospitalized, 44 (10.7%) were transported to other regional hospitals, and 16 (3.9%) were discharged to home in quarantine.

Chest X-ray was performed in 128 (31.1%) of 411 patients and was reported as suggestive of interstitial pneumonia in 79 (61.7%) patients and negative in 49 (38.3%). Chest CT was performed in 317 (77.2%) of 411 patients and showed interstitial pneumonia in 304 (96%), whereas was negative in 13 (4%). It showed pleural effusion in 29 (9.1%) patients, lymphadenopathy in 37 (11.7%), and pericardial effusion in 27 (8.5%). Of the 38 (9.2%) patients who underwent both the exams, 31 (81.6%) had concordant positive report, 6 (15.8%) had positive CT scan and negative X-Ray report, and 1 (2.6%) had negative CT scan and positive X-Ray report (table 2).

**Table 2.** Variables predicting the events included in the primary outcomes (CPAP/NIV, ICU and death) SARS-CoV-2 patients admitted to the Hospital of Crema between February 21 and March 13, 2020, and followed up until March 19, 2020. X-ray and CT are intended as the first exam performed and percentage in brackets refers to the number of patients who have done the exams. Sums do not add up to the total because of some missing values. In bold Bonferroni adjusted significant p-values (i.e. p-values<0.017).

|  | All patients<br>(411) | Deaths<br>(72) | p value | ICU<br>(28) | p value | CPAP/NIV<br>(112) | p value |
| --- | --- | --- | --- | --- | --- | --- | --- |
| **Age** |  |  |  |  |  |  |  |
| mean (standard deviation) | 66.8 (16.4) | 81.1 (7.5) | **<0.001** | 65.7 (12.1) | 0.24 | 70.9 (10.9) | 0.03 |
| median (range) | 70.5 (1-99) | 80.7 (63-99) |  | 69.1 (33-85) |  | 72.1 (33-91) |  |
| **Sex, n (%)** |  |  |  |  |  |  |  |
| Male | 359 (66.6) | 53 (73.6) | 0.23 | 26 (92.9) | **0.003** | 91 (81.3) | **<0.001** |
| Female | 180 (33.4) | 19 (26.4) |  | 2 (7.1) |  | 21 (18.7) |  |
| **Cough, n (%)** | 151 (36.7) | 8 (11.1) | **<0.001** | 11 (39.3) | 0.77 | 33 (29.5) | 0.06 |
| **Dyspnoea, n (%)** | 131 (31.9) | 39 (54.2) | **<0.001** | 22 (78.6) | **<0.001** | 67 (59.8) | **<0.001** |
| **Any comorbidity, n (%)** | 256 (62.3) | 61 (84.7) | **<0.001** | 23 (82.1) | **0.03** | 87 (77.7) | **<0.001** |
| Hypertension | 193 (47.0) | 48 (66.7) | **<0.001** | 20 (71.4) | **0.007** | 67 (59.8) | **0.001** |
| Cardiovascular diseases | 93 (22.6) | 28 (38.9) | **<0.001** | 9 (32.1) | 0.21 | 37 (33.0) | **0.002** |
| Diabetes | 67 (16.3) | 25 (34.7) | **<0.001** | 7 (25.0) | 0.20 | 33 (29.5) | **<0.001** |
| Pulmonary diseases | 48 (11.7) | 10 (13.9) | 0.52 | 5 (17.9) | 0.29 | 18 (16.1) | 0.09 |
| Renal insufficiency | 22 (5.3) | 11 (15.3) | **<0.001** | 5 (17.9) | **0.002** | 11 (9.8) | **0.014** |
| Malignancies | 33 (8.0) | 9 (12.5) | 0.12 | 2 (7.1) | 0.86 | 8 (7.1) | 0.69 |
| **Number of drugs, n (%)** |  |  |  |  |  |  |  |
| 0 | 124 (33.9) | 10 (15.9) | **<0.001** | 4 (16.0) | 0.03 | 19 (19.0) | **<0.001** |
| <3 | 89 (24.3) | 8 (12.7) |  | 5 (20.0) |  | 20 (20.0) |  |
| 4-6 | 74 (20.2) | 14 (22.2) |  | 5 (20.0) |  | 25 (25.0) |  |
| >7 | 79 (21.6) | 31 (49.2) |  | 11 (44.0) |  | 36 (36.0) |  |
| **ACE inhibitors, n (%)** | 50 (12.2) | 11 (15.3) | 0.37 | 4 (14.3) | 0.72 | 20 (17.9) | 0.03 |
| **Sartans, n (%)** | 60 (14.6) | 14 (19.4) | 0.20 | 9 (32.1) | **0.006** | 22 (19.6) | 0.08 |
| **X-Ray, n (%)** |  |  |  |  |  |  |  |
| probable/possible | 79 (61.7) | 25 (34.7) | **<0.001** | 11 (39.3) | **0.013** | 28 (25.0) | **0.003** |
| negative | 49 (38.3) | 3 (4.2) |  | 1 (3.6) |  | 4 (3.6) |  |
| not done | 283 (68.9) | 44 (61.1) |  | 16 (57.1) |  | 80 (71.4) |  |
| **CT scan, n (%)** |  |  |  |  |  |  |  |
| positive | 304 (96.0) | 50 (100) | 0.07 | 23 (82.1) | 0.46 | 92 (82.1) | 0.019 |
| negative | 13 (4.0) | 0 (0.0) |  | 0 (0.0) |  | 0 (0.0) |  |
| not done | 94 (22.8) | 22 (30.6) |  | 5 (17.9) |  | 20 (17.9) |  |

Laboratory exams at admission showed white blood cell count below 10×10^9^/L in 82% of patients, neutrophil count below 10×10^9^/L in 83.4%, and lymphocyte count below 1×10^9^/L in 55.6%. C-reactive protein, aspartate aminotransferase and lactate dehydrogenase values were elevated in most patients, particularly among those with worst clinical picture and outcome (table 3).

**Table 3.**
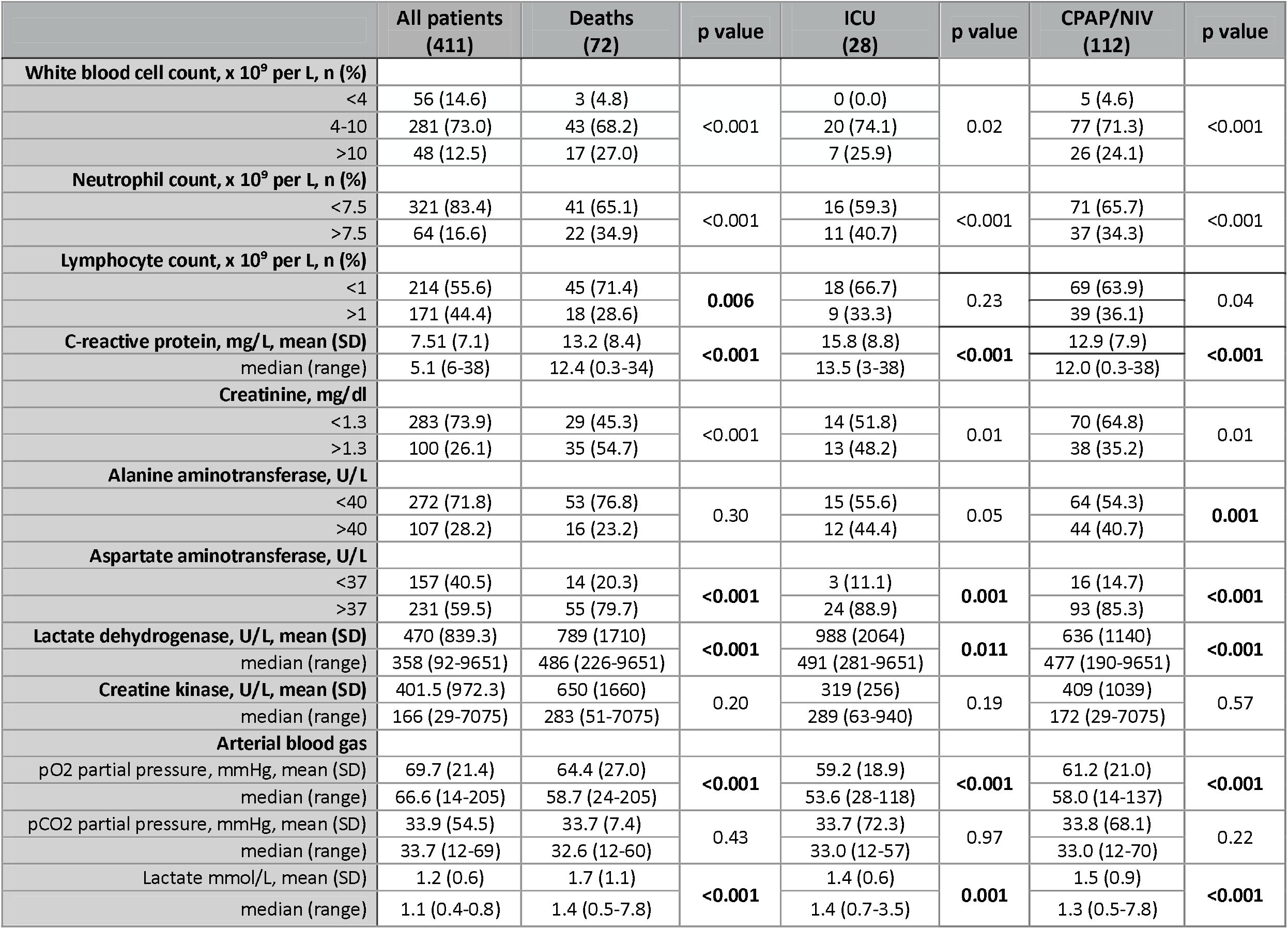
Laboratory findings predicting the events included in the primary outcomes (CPAP/NIV, ICU and death) in SARS-CoV-2 patients admitted to the Hospital of Crema between February 21^st^ and March 13^rd^, 2020, and followed up until March 19, 2020. Sums do not add up to the total because of some missing values. In bold, p values of clinically significant abnormal findings with Bonferroni adjusted significant p- values (i.e. p-values<0.017).

|  | All patients<br>(411) | Deaths<br>(72) | p value | ICU<br>(28) | p value | CPAP/NIV<br>(112) | p value |
| --- | --- | --- | --- | --- | --- | --- | --- |
| **White blood cell count, x 10<sup>9</sup> per L, n (%)** |  |  |  |  |  |  |  |
| <4 | 56 (14.6) | 3 (4.8) | <0.001 | 0 (0.0) | 0.02 | 5 (4.6) | <0.001 |
| 4-10 | 281 (73.0) | 43 (68.2) |  | 20 (74.1) |  | 77 (71.3) |  |
| >10 | 48 (12.5) | 17 (27.0) |  | 7 (25.9) |  | 26 (24.1) |  |
| **Neutrophil count, x 10<sup>9</sup> per L, n (%)** |  |  |  |  |  |  |  |
| <7.5 | 321 (83.4) | 41 (65.1) | <0.001 | 16 (59.3) | <0.001 | 71 (65.7) | <0.001 |
| >7.5 | 64 (16.6) | 22 (34.9) |  | 11 (40.7) |  | 37 (34.3) |  |
| **Lymphocyte count, x 10<sup>9</sup> per L, n (%)** |  |  |  |  |  |  |  |
| <1 | 214 (55.6) | 45 (71.4) | **0.006** | 18 (66.7) | 0.23 | 69 (63.9) | 0.04 |
| >1 | 171 (44.4) | 18 (28.6) |  | 9 (33.3) |  | 39 (36.1) |  |
| **C-reactive protein, mg/L, mean (SD)** | 7.51 (7.1) | 13.2 (8.4) | <0.001 | 15.8 (8.8) | <0.001 | 12.9 (7.9) | <0.001 |
| median (range) | 5.1 (6-38) | 12.4 (0.3-34) |  | 13.5 (3-38) |  | 12.0 (0.3-38) |  |
| **Creatinine, mg/dl** |  |  |  |  |  |  |  |
| <1.3 | 283 (73.9) | 29 (45.3) | <0.001 | 14 (51.8) | 0.01 | 70 (64.8) | 0.01 |
| >1.3 | 100 (26.1) | 35 (54.7) |  | 13 (48.2) |  | 38 (35.2) |  |
| **Alanine aminotransferase, U/L** |  |  |  |  |  |  |  |
| <40 | 272 (71.8) | 53 (76.8) | 0.30 | 15 (55.6) | 0.05 | 64 (54.3) | **0.001** |
| >40 | 107 (28.2) | 16 (23.2) |  | 12 (44.4) |  | 44 (40.7) |  |
| **Aspartate aminotransferase, U/L** |  |  |  |  |  |  |  |
| <37 | 157 (40.5) | 14 (20.3) | <0.001 | 3 (11.1) | **0.001** | 16 (14.7) | <0.001 |
| >37 | 231 (59.5) | 55 (79.7) |  | 24 (88.9) |  | 93 (85.3) |  |
| **Lactate dehydrogenase, U/L, mean (SD)** | 470 (839.3) | 789 (1710) | <0.001 | 988 (2064) | **0.011** | 636 (1140) | <0.001 |
| median (range) | 358 (92-9651) | 486 (226-9651) |  | 491 (281-9651) |  | 477 (190-9651) |  |
| **Creatine kinase, U/L, mean (SD)** | 401.5 (972.3) | 650 (1660) | 0.20 | 319 (256) | 0.19 | 409 (1039) | 0.57 |
| median (range) | 166 (29-7075) | 283 (51-7075) |  | 289 (63-940) |  | 172 (29-7075) |  |
| **Arterial blood gas** |  |  |  |  |  |  |  |
| pO2 partial pressure, mmHg, mean (SD) | 69.7 (21.4) | 64.4 (27.0) | <0.001 | 59.2 (18.9) | <0.001 | 61.2 (21.0) | <0.001 |
| median (range) | 66.6 (14-205) | 58.7 (24-205) |  | 53.6 (28-118) |  | 58.0 (14-137) |  |
| pCO2 partial pressure, mmHg, mean (SD) | 33.9 (54.5) | 33.7 (7.4) | 0.43 | 33.7 (72.3) | 0.97 | 33.8 (68.1) | 0.22 |
| median (range) | 33.7 (12-69) | 32.6 (12-60) |  | 33.0 (12-57) |  | 33.0 (12-70) |  |
| Lactate mmol/L, mean (SD) | 1.2 (0.6) | 1.7 (1.1) | <0.001 | 1.4 (0.6) | **0.001** | 1.5 (0.9) | <0.001 |
| median (range) | 1.1 (0.4-0.8) | 1.4 (0.5-7.8) |  | 1.4 (0.7-3.5) |  | 1.3 (0.5-7.8) |  |

Six patients died within the first 72 hours and another 66 patients within two weeks after hospitalization, giving an overall fatality rate of 17.5%. No death occurred in patients aged below 60 years. In the older age groups, the number of deaths was 26 (88.5% males) in the decade 70-79 and 31 (71% males) in the decade 80-89 (fig. 2). Non-invasive respiratory support was needed in 112 (27.2%) patients, with 28 (6.8%) requiring ICU admission.

**Figure 2.**
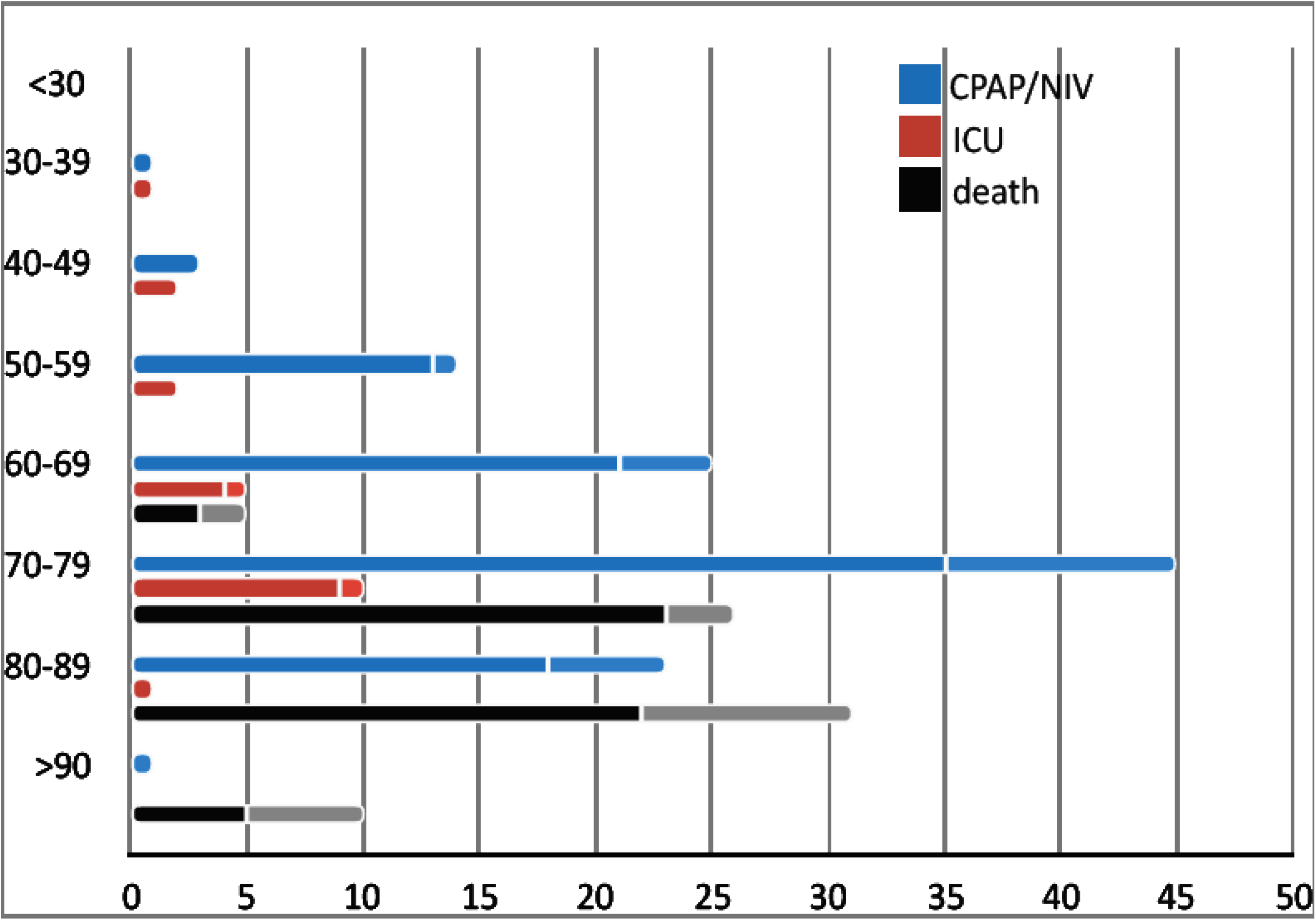
Age and sex-stratification of the outcomes (CPAP/NIV, ICU admission, and death) in 411 SARS-CoV-2 patients. Bars are number of patients. Females are represented in the upper softer bars.

Antiretroviral treatment with ritonavir plus lopinavir was started within the first 72 hours in 42.3% of patients, in combination with hydroxychloroquine sulfate in 63% of them.

### Course, outcome and follow-up in nasopharyngeal swab negative patients

Among the 128 patients negative at nasopharyngeal swab, the median age was 67.7 years (range 1-98), with a preponderance of males (63.3%). Thirty-six (28.1%) patients arrived at hospital with an ambulance. One 36 year-old woman with recurring epileptic seizures and possible meningoencephalitis was moved to another hospital. Seven patients (78 to 97 years) died. Chest CT could be performed in 5 patients and it was positive in all. Of 128 patients, 82 (64%) were hospitalized and 25 (19.5%) were discharged to home in quarantine. None was moved to other regional hospitals.

Chest X-ray was performed in 59 (46%) of 128 patients and was reported as suggestive of interstitial pneumonia in 30 (50.8%) and negative in 29 (49.2%). CT scan was performed in 87 (68%) of 128 patients and showed interstitial pneumonia in 76 (87.3%), whereas was negative in 11 (12.7%). It showed pleural effusion in 6 (6.9%) patients, lymphadenopathy in 8 (9.2%), and pericardial effusion in 41 (47.1%). Of the 21 (16.4%) patients who underwent both the exams, 16 (76.2%) had concordant positive report, 3 (14.3%) had positive CT scan and negative X-Ray report, and 2 (9.5%) had concordant negative report.

Laboratory exams at admission showed white blood cell count below 10×10^9^/L in 61% of patients, neutrophil count below 10×10^9^/L in 67.4%, and lymphocyte count below 1×10^9^/L in 32.2%. C-reactive protein, aspartate aminotransferase and lactate dehydrogenase values were mildly elevated in most patients, particularly those with worst clinical picture and outcome.

Two patients died within the first 72 hours and another 15 patients within two weeks after hospitalization, giving an overall fatality rate of 13.3%. Nine (53%) of the 17 patients who died had positive CT scan as compared to 25 (22.5%) in those hospitalized or discharged. Non-invasive respiratory support was needed in 22 (17.2%) patients, with 4 (3.1%) requiring ICU admission.

Antiretroviral treatment with ritonavir plus lopinavir was started in 27 (42.3%) patients, in combination with hydroxychloroquine sulfate in 82.1% of them.

Of 128 patients, 32 (25%) repeated nasopharyngeal swab (median time 8 days, range 1-26); 13 (40.6%) patients turned positive and 8 of them had either X-ray or CT positive. None was admitted to ICU and 3 died during hospitalization.

### Diagnostic accuracy of nasopharyngeal swab

Because a number of patients had clinical and chest CT findings indicative of interstitial pneumonia, irrespective of the result of the nasopharyngeal swab, we calculated its diagnostic accuracy. To this aim, we used the report of chest CT as gold standard for the diagnosis of interstitial pneumonia and calculated the diagnostic accuracy of the first nasopharyngeal swab in entire study population of 539 patients. The sensitivity was 0.80 (CI 0.76-0.84), the specificity was 0.45 (CI 0.25-0.65), the positive predicting value was 0.96, and the negative predictive value was 0.13.

### Comorbidity pattern and risk profile for outcome in SARS-CoV-2 patients

Older age, cough and dyspnea at onset, history of hypertension, cardiovascular diseases, diabetes, renal insufficiency, intake of 7 or more drugs and positive chest X-ray at admission were significantly associated with death (table 2). Among laboratory exams at admission, low lymphocyte count, high C-reactive protein, aspartate aminotransferase and lactate dehydrogenase, and low PO_2_ partial pressure with high lactate at arterial blood gas analysis were also significantly associated with death (table 3).

Comorbidity network analysis was performed on the reduced set cohort of SARS-CoV-2 patients born between 1920 and 1980, and suffering for at least one of the diseases included in the analysis. Subsequently, the reduced sample was divided in three age groups: 1920-1940, 1941-1960 and 1961-1980. The comorbidity networks obtained from those groups (fig. 3 a, b and c) showed different topological properties. The community detection algorithm was able to identify two modules only for the age groups 1920-1940 and 1961-1980, whereas only one community characterized the age group 1941-1960. It means that in the two extremes of the range of age there are two different groups of diseases (pink and green in fig. 3 a and c) and the number of patients who share two diseases is more similar between two nodes of the same community than between two nodes of different communities. Conversely, the one community configuration found in the 1941-1960 age group is associated with a homogeneous distribution of the number of patients who share two diseases across the whole network. The different weighted degree (i.e. the node size in fig. 3) highlights a bimodal disease distribution among patients. Specifically, cardiovascular disease (CaD), respiratory diseases (ReD), diabetes and hypertension (HT) seem to be more largely present among patients, along with chronic renal insufficiency (ReI) and cancer in the 1920-1940 group, cancer in the 1941-1960 group and asthma in the 1961-1980 group. Finally, the same set of patients included in the analysis was divided in two groups according to the outcome deceased or survived. Notably, only one module characterized the deceased network, whereas the survived network showed two communities. This means that deceased patients were more homogeneously distributed across diseases than survived patients. The weighted degree distribution showed that deceased patients shared equally six important conditions: CAD, ReD, cancer, HT, diabetes and ReI. At the same time, survived patients show the same set of diseases as the most common across patients except ReI (fig. 3 d and e). Figure 4 shows the test of the statistically significant difference between normalized Laplacian of survived and deceased networks. The randomization approach allowed the rejection of the null hypothesis (the two graphs have the same structure) with p < 0.007.

**Figure 3.**
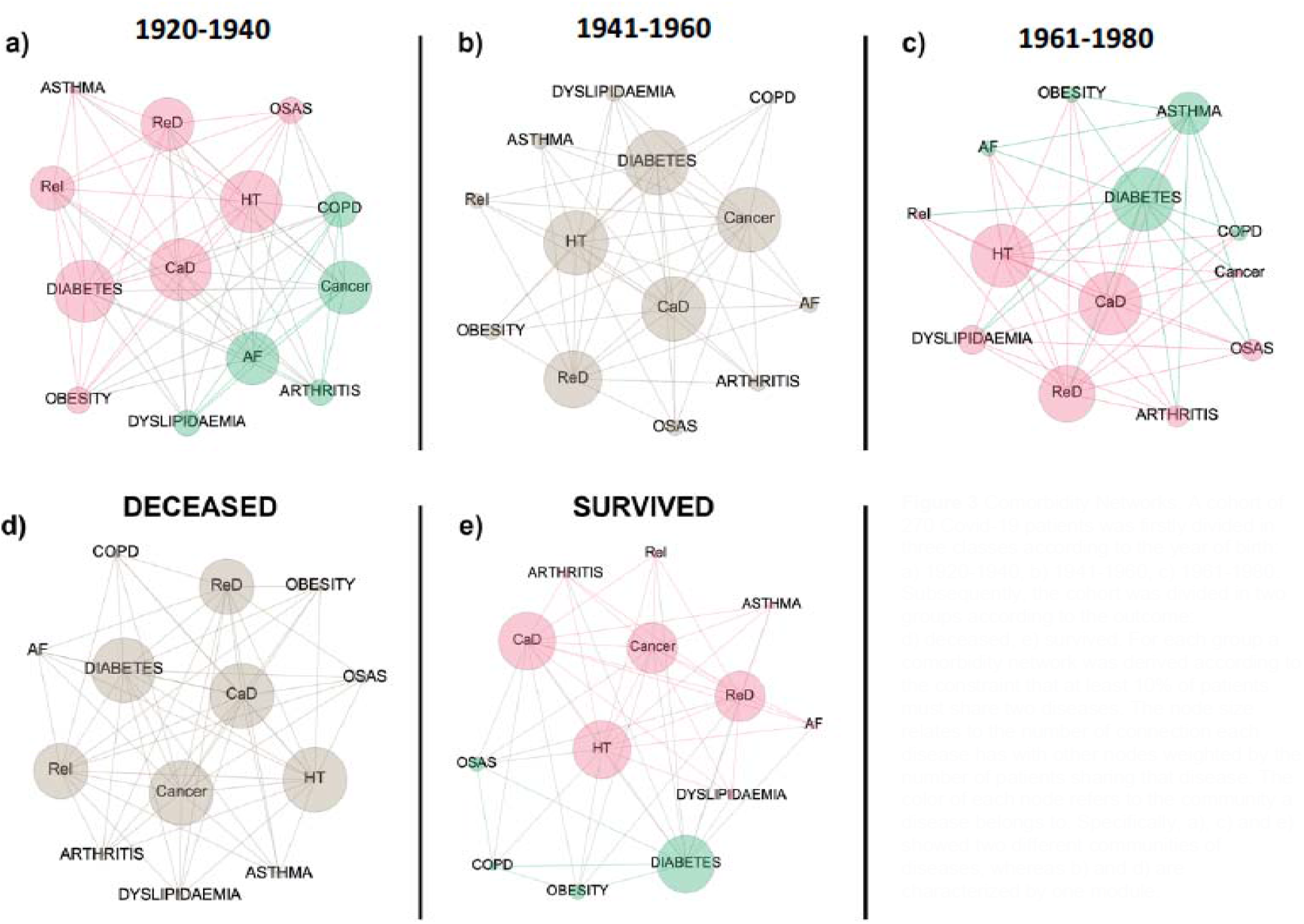
Comorbidity Networks. A cohort of 270 Covid-19 patients was firstly divided in three classes according to the year of birth: a) 1920-1940, b) 1941-1960, c) 1961-1980. Subsequently, the cohort was divided in two groups according to the outcome: d) deceased, e) survived. For each group a comorbidity network was derived according to the constraint that at least 10% of patients must share two diseases. The node size relates to the number of connection each disease has with other nodes weighted by the number of patients sharing that disease. The color of each node refers to the community a disease belongs to. Specifically, a), c) and e) showed two different communities of diseases, whereas b) and d) are characterized by one module.

**Figure 4.**
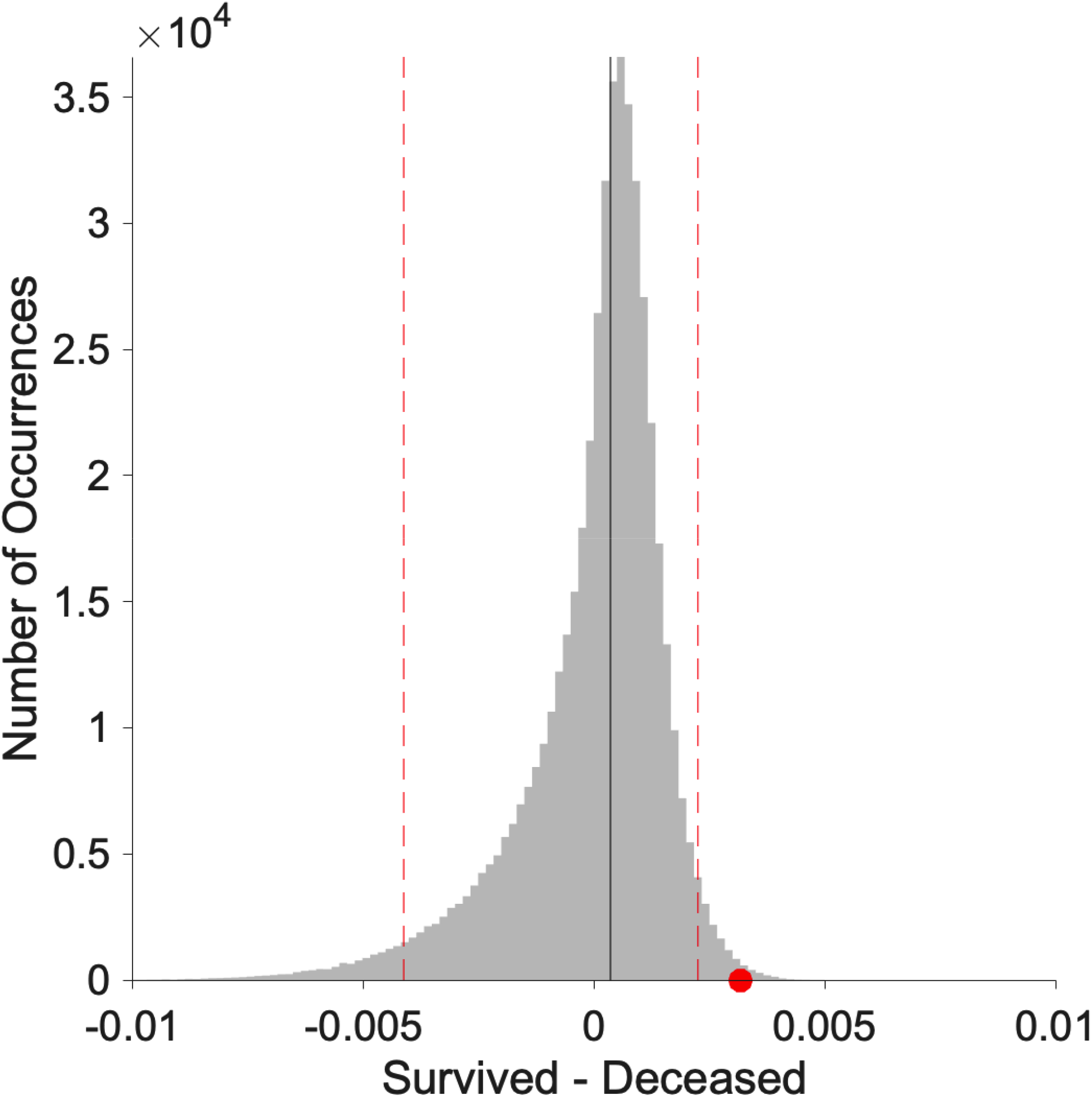
Statistically significant difference between normalized Laplacian of deceased and survived networks. The difference resulted significant with p<0.007. The grey distribution is the result of the difference between 500,000 randomization of survived and deceased normalized Laplacian matrices. The red dot shows the difference between the original survived and deceased normalized Laplacian matrices. The dotted vertical lines represent the 0.05 tails of the distribution.

## Discussion

We reported the first systematic analysis on hospitalized SARS-CoV-2 patients soon after the outbreak in Italy. The strength of our study was the availability of homogenous data from all consecutive patients admitted to a public hospital and suspected to harbor SARS-CoV-2 infection. Therefore, we could investigate the course of any patients presenting with symptoms indicative of possible interstitial pneumonia, including those who resulted negative at nasopharyngeal swab. One limitation is the report of the events occurring soon after the outbreak, during a period in which many information on the disease course were not yet available, and with a hospital-based perspective.

We examined 411 patients with laboratory-confirmed SARS-CoV-2 at nasopharyngeal swab and compared their clinical features and course to that of 128 swab negative patients. Overall, the percentage of SARS-CoV-2 patients that arrived at hospital with an ambulance and required non-invasive respiratory support or ICU admittance was as twice as that of swab negative patients. Nevertheless, the fatality rate was only about 4% higher in SARS-CoV-2 patients. At the same time, chest CT was reported as positive in nearly 60% of swab negative patients, as previously reported,^9^ and mainly among those who died. These findings indicate that a number of swab negative patients were admitted to the hospital because of an interstitial pneumonia, which could be attributed to SARS-CoV-2 in 13 of the 32 patients in whom it was possible to perform the follow-up nasopharyngeal swab that resulted positive.

Even though no study has assessed the diagnostic accuracy of RT-PCR on nasopharyngeal swab in suspected SARS-CoV-2 patients, it has been reported ^10,11^ that it could be negative in patients eventually receiving this diagnosis. Because nasopharyngeal swab is currently used as screening tools in the population, its sensitivity value, namely the percentage of true positive, is important. We sought to address this issue by using chest CT as the gold standard for the diagnosis of interstitial pneumonia on the entire study population of 539 patients. In our cohort, we estimated a sensitivity of 80%. This result was based on the analysis of severely symptomatic patients, thus might not be widely applicable. Nevertheless, it provided an estimation that could be useful for hospital-based surveys.

In our cohort, SARS-CoV-2 predominantly affected males. Our rate of 66.6% males confirmed those reported in China ranging between 54.3% and 73%.^1-3,12-15^ Only one study^16^ showed an inverse trend with 56% females affected. No biological explanation for this unexpected gender distribution is currently available.

Fatality rate related to the whole population of SARS-CoV-2 subjects is unreliable and of limited usefulness in the pandemic, while urgent strategies are needed to improve patients’ care and healthcare system capability. Indeed, the number of SARS-CoV-2 subjects experiencing mild, or non-respiratory symptoms (e.g. gastrointestinal), plus those asymptomatic are unknown because of the variability in testing policies. Since hospitalized patients carry the highest risk of dismal outcome, the inherent fatality rate should be used also to compare findings across countries. Our hospital-based fatality rate in SARS-CoV-2 patients was 17.5%. Previous Chinese studies on hospitalized patients reported rates of 28% in 191 patients^13^ and 21.8% in 201 patients.^15^ Three studies including 99,^1^ 137,^16^ and 41^2^ patients reported lower rates ranging between 11%^1,16^ and 14.6%.^2^ Another two studies reported much lower rates of 1.4% and 4.3%.^3^ We did not record any death below 60 years. Mortality increased across the decades and it was 6.6% in 60-69 years old, 21.1% in 70-79 years old, 38.8% in 80-89 years old and 83.3% above 90 years. Nearly 74% of the deaths occurred in males, which may be not surprising based on the predominantly male distribution of the diseases. Nevertheless, the fatality rate among males exceeded by 10% that of the whole population of SARS-CoV-2 patients, suggesting that the disease could more severely affect males. A similar difference between deaths and infected was reported in one ^13^ out of three previous studies.^14,15^ One only study provided age-stratified fatality rates of patients admitted to ICU ^17^, thus further studies are warranted to confirm our findings on hospitalized patients.

Non-invasive respiratory support was needed in 27% of SARS-CoV-2 patients and ICU admission rate was 6.8%. Previous Chinese studies^1-3,16^ reported ICU admission rates between 4.3% and 31.7%, with fatality rate between 11% and 14.6%. A recent Italian study focused on ICU patients ^17^ reported overall fatality rate of 26%. This comparison may suggest that a larger availability of ICU beds might be associated with lower fatality rates. To investigate this hypothesis, we analyzed the distribution of the fatality rates in the decades 60-69 (76 patients) and 70-79 (123 patients) (fig. 2). The proportion of patients in CPAP/NIV and ICU (32.9% and 6.5% in 60-69, respectively and 36.5% and 8.1% in 70-79, respectively) did not differ. Nevertheless, the fatality rate was 30% higher in the 70-79 (47%; 21/45; p=0.02) than 60-69 decade (16%; 4/25). Among patients admitted to ICU, the fatality rate was 40% (4/10) in the 70-79 years old, the same finding reported in the Italian larger series ^17^.

Tentative explanation of this difference could be the generic higher risk related to aging, because patients in the two groups of age seemed to not significantly differ in terms of distribution of comorbidities and use of more than 4 drug (10% and 5% higher in 70-79 years old, respectively). Thus, even though it might be possible that higher ICU admission rate could have reduced the mortality in the older age group, leading to a lower overall fatality rate, older age per se might also play a major role on dismal outcome.

All previous studied ^1-3,12-16,18,19^ reported high frequency of comorbidities mainly among patients with poorer outcome, suggesting a role in enhancing SARS-CoV-2 morbidity.^20^ To better correlate relationship and frequency of comorbidities to age and outcome, we adopted a network analysis approach. Our findings showed that in deceased patients, six conditions with intrinsic potentiality to induce multi-organ involvement (i.e. CAD, ReD, cancer, HT, diabetes and ReI) were equally weighted as compared to surviving patients. The same homogeneous distribution was found in the age group between 60 and 80 years, in which deaths were 43% (31/72) of the total recorded. Even though indirectly and in a relatively small sample size, these findings seem suggesting that SARS-CoV-2 morbidity is enhanced, and possibly even triggered by pre-existing multi-organ impairment.^20^

Finally, a major concern was raised on the use ACE inhibitors and sartans because SARS-CoV-2 uses the ACE2 (angiotensin-converting enzyme 2) protein as the receptor to entry the cells.^21^ Our results demonstrated that these classes of drug did not have any influence on patients’ outcome.

## Data Availability

The data that support the findings of this study are available from the corresponding author.

## Acknowledgements

We are deeply indebted with all the nurses of the Hospital of Crema, whose passion, skills and fortitude in taking care of patients was a bastion to limit the wave of suffering that in a few weeks have overwhelmed an entire community. We are particularly grateful to Dr. Maria Lidia Sinatra, Health Management Unit and Dr. Anna Maria Bona, Nurse Management Unit. We also indebted with the staff of all the departments, for the support provided to the study.

We also wish to acknowledge our collaborators at the RADIOLOGY UNIT (Drs. Acqualagna Renato, Astorri Silvia, Bonardi Roberto, Borghetti Maurizio, Conti Maria Paola, Garavaglia Marco, Gandolfi Silvia, Maldotti Marco, Limonta Simone, Parziale Maurizio Giuseppe, Spinazzola Angelo, Padrenostro Mauro); GASTROENTEROLOGY UNIT (Drs. Alicante Saverio, Brambilla Gianfranco, Iiritano Elena, Londoni Claudio, Manfredi Guido, Menozzi Fernanda, Romeo Samanta, Pedaci Marianna EMERGENCY MEDICINE UNIT Accordino Silvia, Cacco Silvia, De Matthaeis Anna, Formagnana Pietro, La Boria Elisa, Masotti Michela, Provini Stella, Seresini Monica, Severgnini Silvia, Taverni Silvana, Valsecchi Cesare, Vezza Carlotta); PNEUMOLOGY UNIT 1 (Drs. Luca Bilucaglia, Michele Ceruti, Alessia Edallo, Fabrizio Mauri, Mara Parati, Emanuele Rana, Simona Scorsetti, Massimiliano Villani); PNEUMOLOGY UNIT 2 (Drs. Besozzi Alessandra, Chiesa Lodovico, Giunti Valeria); INTENSIVE CARE UNIT (Drs. Bianchi Fulgenzia, Biazzi Antonella, Bisicchia Maria Cristina, Bonfanti Giuseppina, Caruso Andrea, Cerisara Marco, Chiarenza Federica, Foglio Elena, Francavilla Nicola, Gigliotti Alberto, Meglio Giuseppina, Perotti Vittorio, Adriani Luca, Bergamaschini Giulio, Borromeo Raffaella, Comassi Paolo, Corallo Sandro, Lupi Giuseppe, Troiano Carmine, Villani Pier Giorgio); EMERGENCY UNIT (Drs. Guerra Claudia, Piantelli Miriam, Susca Micaela, Galmozzi Attilio, Zuccon William, Sportelli Silvia, Corti Serena, Depetri Federica, Ruini Alain, Kostihova Anna, Dolci Francesco, Bertin Elena)

